# Task-specific changes in representation and connectivity between parietal and occipito-temporal areas following left hemisphere stroke

**DOI:** 10.1101/2025.01.26.25321127

**Authors:** Zuo Zhang, Ajay Halai, Stefania Bracci, Gloria Pizzamiglio, Matthew Lambon Ralph, Elisabeth Rounis

**Author notes:** Corresponding authors: Dr Elisabeth Rounis, Neurology Department, OPD8 West Middlesex University Hospital, Chelsea and Westminster NHS Foundation Trust, London TW7 6AF, Professor Matthew Lambon Ralph, MRC CBU, University of Cambridge, 15 Chaucer Road, CB2 7 EF.

## Abstract

Recent studies highlight a critical role of the human lateral occipitotemporal cortex (LOTC) in action understanding, particularly in how we perceive and interpret movements and interactions with tools and bodies. This region, along with connected fronto-parietal areas, has been implicated in limb apraxia, a disorder that affects skilled movement after stroke.

In this study, we focused on whether there were any changes in brain activity or connectivity in areas involving the LOTC and its interconnected perceptuo-motor network after left hemisphere stroke, a common cause of limb apraxia.

We recruited 29 stroke patients and 19 age-matched healthy participants. Using functional neuroimaging, we asked participants to observe static photographs of familiar tools, bodies, non-tool objects, and scrambled images while performing a simple attention task (1-back task). The goal was to map the brain areas specifically involved in tool-related and body-related visual processing. Whole-brain analysis revealed activity changes related to tool and body stimuli in the LOTC and fronto-parietal regions. Interestingly, there were no significant differences in activation between stroke patients and controls based on the overall whole-brain analysis. However, deeper analyses revealed critical group-related differences. Using representational similarity analysis (RSA), we found that stroke patients showed reduced ability to discriminate between tools and non-tools within the right LOTC. Psychophysiological interactions (PPI) analysis further indicated increased connectivity between the left inferior parietal sulcus (IPS) and the left LOTC in stroke patients during tool-related tasks, a finding that correlated with their performance on meaningless imitation apraxia task.

Our findings suggest that limb apraxia after stroke is associated with altered representation of tools and functional connectivity in the perceptuo-motor network, particularly involving parietal and occipitotemporal regions. These results suggest perceptual deficits may be more relevant than previously reported in limb apraxia after stroke.

**Highlights:**

- Consistent activations in the lateral occipitotemporal cortex (LOTC) were identified during tool and body localizer tasks in both stroke patients and healthy controls.
- Representational similarity analysis (RSA) revealed reduced discrimination between tools and non-tools in the right LOTC of stroke patients.
- Differential task-related functional connectivity was observed between the LOTC and intraparietal sulcus (IPS) regions in stroke patients compared to controls during the tools localizer task.

These findings provide insights into altered neural representations and connectivity underlying limb apraxia following left hemisphere stroke.

## Introduction

Limb apraxia (thereafter referred to as ‘apraxia’) is a disabling condition that commonly follows a stroke, impairing patients’ ability to perform everyday tasks, such as making a cup of tea, or gesturing. Traditionally, research has focused on lesion-symptom mapping to identify the brain regions involved in this disorder, often within the framework of the perception-action model (Buxbaum et al., 2014, Hoeren et al., 2014). Although these studies and more recent investigations into structural disconnections of white matter (Rounis et al., 2024) have provided valuable insights, they often overlook how dynamic functional connectivity within perceptuo-motor networks contributes to the behavioural deficits observed in apraxia. This study represents an important advancement by employing task-based functional MRI (fMRI) to explore how brain networks interact during the perception of bodies and tools, providing a more comprehensive view than lesion-symptom mapping alone. By probing disruptions in neural information-tuning and connectivity, fMRI offers deeper insights into the neural mechanisms underlying apraxia (Stefaniak et al., 2020).

Apraxia has traditionally been classified into two main categories: ‘conceptual’ and ‘production’ deficits, each associated with distinct action pathways. The ‘indirect’ pathway involves activating action sequences from memory, which when damaged leads to ideational apraxia. The ‘direct’ pathway, which relies on visual analysis to reproduce movements, is linked to ideomotor apraxia. Whereas the former is usually identified by asking patients to demonstrate familiar gestures or use of tools, the latter is demonstrated by asking them to imitate meaningless gestures. According to the perception-action model, these deficits correspond to distinct brain pathways: the ventro-dorsal stream for ideational deficits and the dorso-dorsal visuomotor stream for ideomotor deficits (Goodale and Milner, 1992;Binkofski and Buxbaum, 2013).

However, this dual-pathway model does not fully account for the wide range of behavioural deficits in apraxia (Buxbaum and Randerath, 2018; Rounis and Binkofski, 2023). One reason for this is that tasks used to assess conceptual and production deficits often involve imprecise and subjective measures (they often rely on experts viewing patients doing gestures and scoring whether they were done correctly or not). Moreover, these tasks may engage overlapping cognitive processes, blurring the distinction between them. Recent data driven techniques have capitalised on the patient variability in performance on apraxia tasks to capture distinct cognitive processes underlying them (Rounis et al., 2021; Schmidt et al., 2022).

There is growing evidence that the functional and anatomical separation of the dual-stream hypothesis is not as clear-cut as previously thought with areas between the two stream often being implicated in several of deficits (Cloutman, 2013; Weiller et al., 2011; Hoeren et al., 2014;Goldenberg and Karnath, 2006; Kalénine et al., 2010). Anatomical connections between these streams, identified through diffusion tractography, suggest significant integration between them (Catani and ffytche, 2005; Ramayya et al., 2010; Heilman and Watson, 2008; Umarova et al., 2010; Rounis et al., 2024). This complexity points to the need for more nuanced models to explain the diverse manifestations of apraxia.

Of particular interest is the lateral occipitotemporal cortex (LOTC), which lies at the intersection of both streams. Although situated within the ventral stream, research suggests that the LOTC may function as part of the dorsal stream due to its white matter connections (Zimmermann et al., 2018). Lesion-symptom mapping studies have implicated LOTC in both ideomotor and ideational apraxia, linking it to deficits in meaningless gesture imitation (Sperber et al., 2019) (Pizzamiglio et al., 2019b) and gesture recognition in patients with aphasia (Vigliocco et al., 2020). The LOTC, along with the adjacent extrastriate body area (EBA), is responsive to the perception of manipulable objects and body parts (Astafiev et al., 2004; Lingnau and Downing, 2015; Bracci et al., 2018). These regions play a role in motor planning, encoding postural configurations required for actions, particularly in anticipating future movements (Astafiev et al., 2004; Zimmermann et al., 2012; Zhang et al., 2021; Zhang et al., 2023). Both the LOTC and EBA are connected to parietal visuomotor regions, integrating into the dorso-dorsal stream to relay information for action guidance. Their perceptual role has been demonstrated in localizer tasks (Gallivan et al., 2013).

While parietal lesions have been highlighted in previous literature, surprisingly few lesion-symptom mapping studies have identified their specific role in limb apraxia (Goldenberg, 2009). Given the role of the extrastriate body area and LOTC in action planning highlighted above, we hypothesised that apraxic deficits may arise from poor information coding in LOTC and/or from impaired perceptual information relayed from the LOTC, affecting connectivity between this region and fronto-parietal areas in the dorsal stream.

In order to determine whether stroke patients exhibit changes in activity or processing of LOTC areas and their connectivity within the network comprising their interconnected perceptuo-motor regions, we used a perceptual localizer task (Gallivan et al., 2013). We examined the brain’s response to body and tool stimuli in both healthy participants and patients with left hemisphere stroke. This allowed us to investigate processing of tool- and body-stimuli in these areas and to map the functional connectivity between the LOTC and key motor and perceptual areas and compare the results between groups, assessing whether stroke-related disruptions in processing or in functional connectivity within this network correlate with the presence and severity of apraxic deficits. Understanding these changes may help elucidate therapeutic targets for subtypes of apraxia (Ant et al., 2019; Rounis and Binkofski, 2023).

## Material and Methods

### Participants

Twenty-nine righted-handed patients with chronic left hemisphere stroke and nineteen healthy volunteers were recruited to participate in this study. The patient genders comprised 11 females, 18 males; their mean age was 59.5 years, age range = 29-79 years. The healthy volunteer gender distributions were 12 females and 7 males; their mean age was 60.5 years, age range 28-79 years. All participants had normal or corrected-to normal vision and no history of any other neurologic or psychiatric diseases. Full written consent according to the declaration of Helsinki was obtained from all participants. The study was approved by the Health Research Authority, South Central – Berkshire Ethics Committee. Participants attended the Oxford Centre for Magnetic Resonance Imaging at the University of Oxford for neuropsychological testing and functional brain imaging. The study procedures or analyses were not pre-registered prior to the research.

### Neuropsychological assessments

Both healthy participants and stroke patients performed a battery of praxis tasks that was originally developed as part of the Birmingham Cognitive Screen (‘BCOS’) to test apraxia using their unaffected, non-dominant, hand as is standardly done in this apraxia screening (Bickerton et al., 2012). The screening took part on the same day as the fMRI study. Participants were counterbalanced whether they were tested before or after they underwent imaging.

They underwent a ‘gesture production’ task, in which participants were asked to pantomime three transitive gestures demonstrating the use of tools (a hammer, a salt cellar and a glass) and three intransitive gestures demonstrating familiar non-tool related gestures (namely how to hitch hike, perform a military salute and stop). They also underwent a ‘meaningless gesture imitation’ task, in which participants imitated meaningless postures and gestures involving 4 hand and 6 finger gestures (as in (Rounis et al., 2016)). Gestures were videotaped and scored as correct or incorrect using the following scoring system: two points were given for a gesture that was correctly and precisely imitated after the 1st presentation; 1 points if the gesture was correct and precise after the 2nd presentation; 0 point if patients made no response or incorrectly imitated the gesture after the 2nd presentation (e.g., incorrect spatial relationship between hand and head, or incorrect finger/hand position), or showed perseveration from previous item(s) after the 2nd presentation. The total correct score (maximum=12 for the gesture production and 20 for meaningless imitation, both converted out of 100) was used in the analyses.

Two independent coders who were blind to the participants were patients or healthy controls (G.P., E.R.) scored the videos for each participant and each task. The final score for each task consisted of the average between the two scores. The average inter-coder reliability for all recorded tasks, defined by Cohen’s Kappa were: 1) patient group: 0.76 for gesture production, 0.8 for meaningless gesture imitation, (together averaging= 0.78), 2) healthy volunteer group: 0.88 for gesture production, 0.85 for meaningless gesture imitation (averaging 0.86) demonstrating similar inter-rater reliability with previous studies (Buxbaum et al., 2005).

### fMRI Localiser task conditions

Participants performed a category ‘Localiser’ task, already described in Gallivan et al. (2013). Each participant completed one run of the localizer scan that included stimulus blocks of colour photographs projected onto a 2D screen consisting of 4 image types: 1) familiar **tools** (87 different identities), 2) headless **bodies** (87 different identities, 44 of which were females), 3) non-tool objects (87 different identities) and 4) scrambled versions of these same stimuli. Each image subtended 15° of the subject’s visual field. To provide a fixation point, a small black circle was superimposed at the centre of each image.

The whole run lasted 6 minutes 10 seconds and was composed of six blocks for each of the ‘Tools’, ‘Bodies’, and ‘Non Tools’ conditions, seven blocks for the ‘Scrambled’ images, and one fixation/baseline block placed at the beginning and at the end of the run. Stimulus blocks (Tools, Bodies and Non Tools) were organized into sets of three, separated by a scrambled block, balanced for block order within a single run. Stimuli were organized into separate 14.4s blocks, with 18 photos per block, presented at a rate of 400ms per photo with a 400ms inter-stimulus interval.

To encourage participants to maintain attention throughout the localizer scans, subjects performed a one-back task throughout, whereby responses were made, via a button press, whenever two successive photos were identical. The frequency of these repetitions was less than 20% of the trials. Two types of response errors were recorded: false positives and misses. False positives indicate erroneous responses when participants pressed buttons that were not corresponding to a repeating trial. Missed responses indicate that participants did not respond when there was a repeated image. Though this task was not relevant for the perceptual fMRI study, participants were excluded if their error rates were above 3 standard deviations (SD) of the group’s mean.

### Image acquisition

MRI data were acquired on a Siemens 3T Trio MRI scanner with a 32-channel RF head coil at the University of Oxford Centre for Clinical Magnetic Resonance Research (OCMR). Structural T1-weighted MRI images were acquired using the MP-RAGE sequence (repetition time, 2040ms; echo time 4.7ms; field of view 174×192mm^2^; 192 slices; voxel size, 1×1×1mm^3^). Functional images were acquired using a whole brain echo planar imaging (EPI) sequence (repetition time, 2400ms; echo time, 30ms; flip angle, 87 degrees field of view, 192×192mm^2^; 41 slices; voxel size, 3×3×3.5mm^3^).

### Imaging data preprocessing (BIDS and fMRI prep output)

The raw DICOM data was converted into BIDS format using *heudiconv* (v0.12.0) The data was then pre-processed using fMRIPrep 22.0.0 (full detail can be found in the supplementary material 1) (Esteban et al., 2019; Gorgolewski et al., 2011).

Briefly, the following pre-processing steps were performed on the T1: 1) corrected for intensity non-uniformity; 2) skull-stripping; 3) tissue segmentation; and 4) non-linear normalisation to MNI152NLin2009cAsym space (using cost-function masking for the stroke lesion). The following pre-processing steps were performed on the fMRI data: 1) skull stripped reference volume; 2) estimate motion parameters; 3) slice time correction (middle slice); 4) co-registered to T1; and 5) mapping to MNI152NLin2009cAsym space using one transformation step. We estimated a B0 non-uniformity map using fmriprep’s fieldmap-less approach, where a deformation field is produced by co-registering the EPI reference image to the T1 (intensity inverted) by constraining deformations only along the phase-encoding direction and modulated with an average fieldmap template.

### Imaging data Analyses

For each participant, the fMRI time series were subjected to General Linear Model (GLM) analysis by using SPM12 (Friston et al., 1994) implemented on MATLAB 2019b (The MathWorks, Inc., US). Single subject models consisted of four regressors separately describing the main effect for each of the four conditions (‘Bodies’, ‘Tools’, ‘Non Tools’ and ‘Scrambled’). These regressors were convolved with a canonical haemodynamic response function (HRF) without derivative terms. Head motion was accounted for by adding the six head motion parameters as additional ‘nuisance’ regressors (Friston et al., 1994). Slow signal drifts were removed by using a 1/128Hz high-pass filter. Serial correlations were accounted for with an autoregressive AR (1) model.

In order to obtain the activity maps for bodies and tools-related processing, the subject-level contrast images for ‘Bodies’ vs. all other conditions (contrast weights: 3, −1, −1, −1) and ‘Tools’ vs. all other conditions (contrast weights: −1, 3, −1, −1) were obtained and subjected to group-level one-sample t-tests, respectively. Furthermore, we assessed differences between patients and heathy volunteers by using two-sample t-tests at the whole-brain level. We applied cluster-wise family wise error (FWE) correction for multiple comparisons at *p* < 0.05, with a cluster-forming threshold of p<0.001 (uncorrected) across the whole brain. Anatomical locations of the activation were labelled according to the AAL3 atlas (Rolls et al. 2020).

### Region of Interest (ROI)-based RSA analysis

We created regions of interest (ROI) for the LOTC and EBA based on peak voxels reported in the literature for these regions (Gallivan et al., 2013). We converted the Talairach coordinates (−53, −57, − 3 for the left LOTC and −49, −72, 1 for the left EBA) reported in Gallivan et al. (2013) into the MNI coordinates (−56, −57, −10 for the left LOTC and −50, −73, −4 for the EBA) by using the MNI2TAL web tool in the BioImage Suite Web (https://bioimagesuiteweb.github.io/webapp/). Regions of interest were created as 10mm spheres centred at these coordinates. The coordinates for the contralateral right LOTC (56, −57, −10) and right EBA (50, −73, −4) were obtained by mirroring the coordinates from the left hemisphere.

A Representational Similarity Analysis (RSA) was conducted in order to identify differences between patients and controls in categorising tools vs bodies in the localiser tasks (Kriegeskorte et al., 2008) We used the fine-grained sensitivity provided by RSA implemented in the Decoding Toolbox (v3.999F) (Hebart et al., 2014), to calculate the representational distance (i.e., dissimilarity) of the activation patterns between the ‘Tools’ versus ‘Non Tools’ in the LOTC ROIs (or between ‘Bodies vs Non Tools’ in the EBA ROIs) for each participant. The pattern dissimilarity measures were calculated by using 1 – Pearson’s *r* and subjected to the Fisher’s transformation. We tested whether the pattern dissimilarity was significantly different between patients and healthy volunteers for Tools and for Bodies respectively, in the LOTC and EBA ROIs for each hemisphere, described above. For the between-group comparison, a non-parametric one-sided Wilcoxon test was used, because the pattern dissimilarity measures were not normally distributed based on Shapiro-Wilk normality tests (*ps* < 0.01). We hypothesized that the pattern dissimilarity between ‘Tools’ and ‘Non Tools’ (and between Bodies and ‘Non Tools’) was greater in healthy volunteers than in patient, based on the hypothesis that apraxia may lead to reduced perceptual discrimination within task categories, suggested by their inability to discriminate gestures ((Pazzaglia et al., 2008), (Rounis and Binkofski, 2023)).

### Psycho-Physical interactions

Functional connectivity changes between patients and healthy volunteers within the networks engaged in representing ‘Bodies’ and ‘Tools’ were assessed using ‘Psycho-physiological Interactions’ (PPI), a method first described by Friston et al. (1997). The PPI analysis explains responses in one cortical area in terms of an interaction between activity in another cortical area and the influence of an experimental condition. We used this to test the hypothesis that connectivity between the left and right LOTC and EBA may differ between healthy volunteers and stroke patients, during representation of tools and bodies, respectively. This hypothesis is based on previous literature which report LOTC areas to be involved in representing hand postures involved in object use (Gallivan, McLean, Valyear, et al., 2013) and EBA to be involved in hand postures (Zimmermann et al. 2018, Zhang et al. 2021).

We combined the literature-based ROI and individual level LOTC activation to create seed-based ROIs for each participant. To create a LOTC ROI for each participant, we located the peak voxel under the subject-level contrast of ‘Tools’ vs. ‘Scrambled’ within the literature-based 10mm sphere for LOTC, and created another 10mm sphere around this individual-level peak. The intersection between the single participant and literature-based spheres were used as the ROI. To create a ROI for the EBA, the contrast of ‘Bodies’ vs. ‘Scrambled’ was used instead. Three variables were created for the PPI analyses in a generalized linear model: a physiological variable for the BOLD signal in the seed region (LOTC, EBA on the Rt and Lt hemisphere), a psychological variable corresponding to the ‘Tool’ or ‘Bodies’ effect (compared to all the other conditions, as in the main effects’ analyses reported above), respectively, and a psycho-physiological interaction variable.

We extracted BOLD signal from each ROI, adjusted for the effects of the four task conditions. To derive brain interactions at the neuronal level, the BOLD signal was deconvolved through haemodynamic function to the neural level before multiplying with the psychological variable to derive the interaction variable. These three PPI variables (physiological, psychological, and interaction variables) were fed into a GLM analysis, together with six head motion estimates as variables of no interest. Subject-level images for the interaction variable entered in two-sample *t* tests to identify differences between patients and healthy volunteers.

## Results

### Behavioural Results

Table 1 reports the respective patient and healthy volunteer group demographics and performance in the ideomotor apraxia tasks. There was a significant difference in performance between the groups in the meaningless gesture imitation (*t*=−3.65, p=0.002) and gesture production (*t*=−2.35, p=0.03) tasks, respectively. The same was true with regards to performance on the 1-back task localiser. Patients showed significantly higher miss rate (*t*=2.63, *p*=0.012) and false positive rate (*t*=2.22, *p*=0.033) than healthy volunteers. We excluded one participant whose miss rate / false positive rate was outlier by using 3 standard deviation (SD) criteria from further analysis.

### Imaging Results

Two patients were excluded from the analysis because functional MRI data were not available for them (failure of image acquisition due to patient not tolerating the scans).. Another patient was excluded due to high behaviour errors on the 1-back task, mentioned above. The lesion overlay map for the remaining 26 patients is shown in Figure 1. The lesions in this patient sample were located on the left hemisphere, concentrated in the white matter of the frontal cortex, extending to the insula, inferior frontal, precentral and postcentral gyri, and the inferior parietal lobule. The left temporal, occipital, and bilateral cerebellum were also affected, but to a lesser extent.

**Figure 1.**
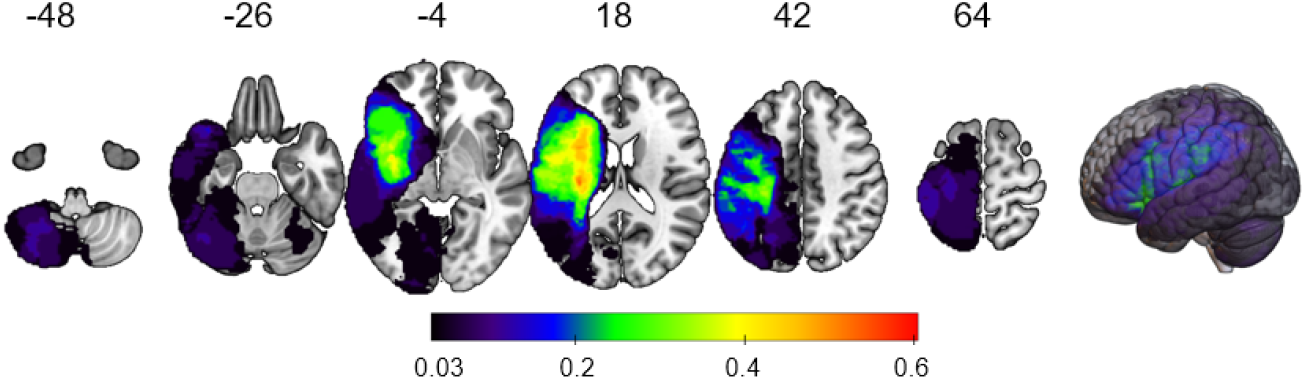
Lesion overlay map. The colour bar indicates the ratio of patients having lesions at a given voxel (1 patient representing a ratio of 0.038). The labels represent the z coordinate in the MNI space.

A group-level analysis investigating effects of our task conditions was performed, combining the patients (N = 26) and healthy volunteers (N = 19) together in the analysis. The overall activations for ‘Tools’, relative to all other conditions are reported in Figure 2A and in Table 2. The overall activations for ‘Bodies’, relative to all other conditions, are reported in Figure 2B and in Table 3. The results reported here were whole-brain corrected at FWE p<0.05, cluster-wise. We conducted between-group comparisons under each experimental condition using the mean activations in each ROI and two-sample *t*-tests (Figure 2C) but did not find any significant differences (*ps* > 0.2).

**Figure 2.**
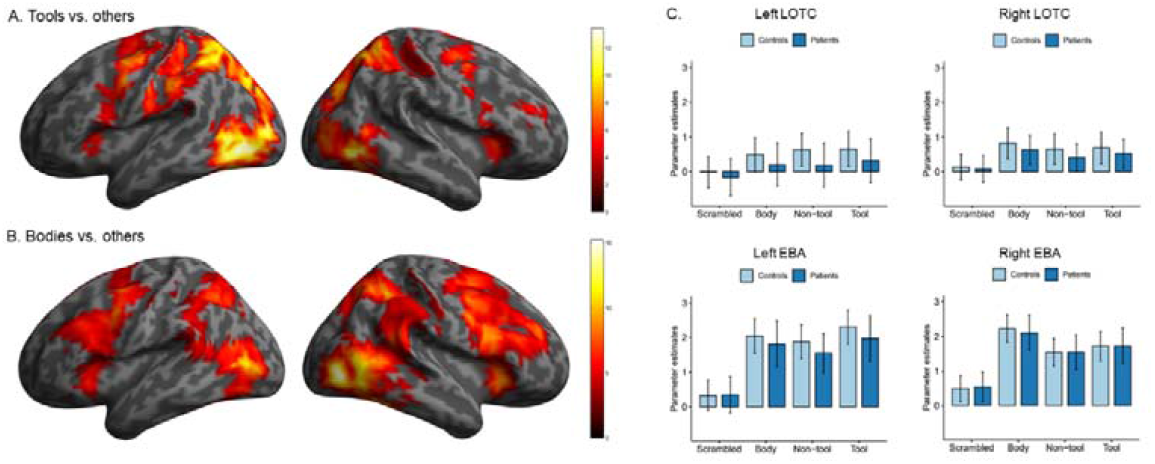
Main effects of Tools and Bodies vs all others in the combined sample of patients and healthy volunteers; This figure represents a projection of voxel-wise statistics on a surface rendering. Colour bar represents *t* statistics. Panel C presents mean activations and 95% confidence intervals in the regions of interest under each experimental condition.

The main effect of Tools vs all the other conditions activated bilateral extrastriate and medial temporal gyri including inferotemporal and lateral occipito-temporal areas, extending to the superior and inferior parietal gyri and dorsal and ventral premotor cortices (Figure 2A). Activity in these areas was greater when viewing Tools than when viewing all other conditions. Though there was a slight predominance of the activations to be left lateralised particularly in the ventral LOTC and dorsal parietal regions, most activations were bilateral. Conversely, activations for Bodies extended from the LOTC to the extrastriate body areas and appeared stronger in the right-hemisphere (but were bilateral as well, Figure 2B). Comparison between patients and healthy volunteers did not reveal significant differences in the activation.

Nevertheless, RSA on the Tools and Bodies based on LOTC and EBA ROIs identified differences for tools and bodies processing, respectively. The pattern of dissimilarity between Tools and Non tools in the right LOTC was greater in healthy volunteers than in patients (Wilcoxon W = 346, *p* = 0.011, 26 patients vs. 19 controls), suggesting that the right LOTC showed higher discrimination between Tools and Non tools in healthy volunteers than in patients (Figure 3). There was also nominally significant difference in the right EBA between Bodies and Non tools, where healthy controls showed higher pattern dissimilarity than patients (Wilcoxon W = 320, *p* = 0.048), though this did not survive correction for multiple comparisons. No significant differences between groups were found in the left LOTC or the left EBA ROIs, even when two patients with lesions at those sites were excluded from the analyses. No patients’ lesions overlapped with the right LOTC/EBA ROIs.

**Figure 3.**
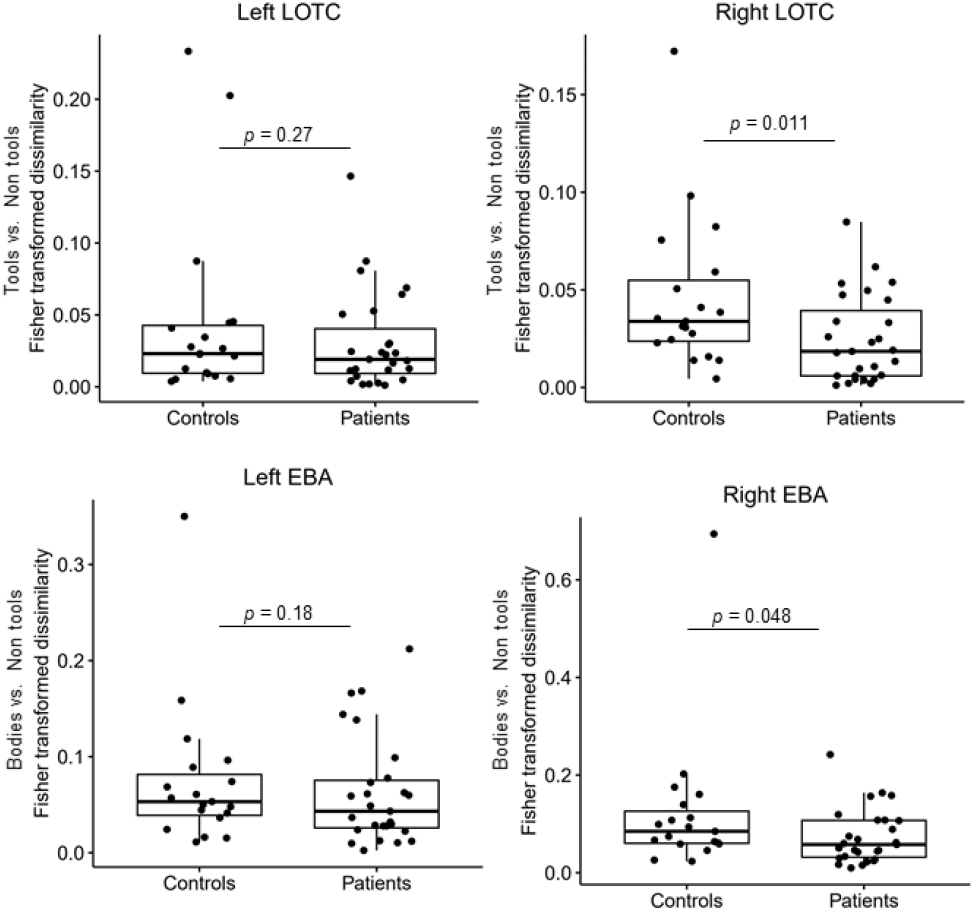
Results for the representational similarity analysis. The y-axis represents the pattern dissimilarity measures (1 - Pearson’s correlation) with Fisher transformation. The *p* value was obtained from one-sided Wilcoxon tests. The top panel showed differences in RSA for Tools vs non Tools for Left and Right LOTC, whereas the bottom panel showed RSA differences for Bodies vs non Tools, respectively.

Finally, we applied PPI analyses to test the hypothesis that the connectivity between LOTC in the patient group differed with the control group under the contrast of Tools vs. all other conditions, and that of EBA under the contrast of Bodies vs. all other conditions. This comparison (26 patients vs. 19 controls) revealed one area whose coupling with the left LOTC was significantly increased in patients compared to healthy volunteers when viewing Tools, namely the left Intraparietal Sulcus (IPS, peak x = −28, y= −76, z= 37, whole brain analysis cluster size = 106 voxels, cluster-level *p*_FWE_ = 0.019, Figure 4). We further considered whether patients with lesions at the seed or result sites might influence our findings, so we repeated the analyses excluding 2 patients with lesions in the LOTC ROI (seed region) and 1 patient with a lesion extending to the left IPS (result region). The results were not significant with whole brain correction (cluster-level *p*_FWE_ =0.061). However this finding survived small volume correction using a 10mm sphere centred at the nearest local maximum to the peak IPS identified from the main effects activation of ‘Tools’ vs ‘Scrambled’ (peak x = −28, y= −79, z= 37, *t* = 4.80, peak-level *p*_FWE_ = 0.001).

**Figure 4.**
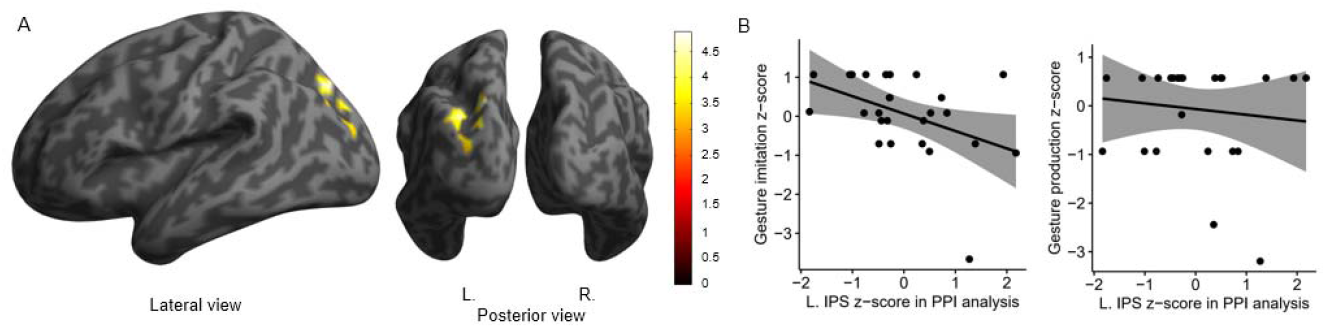
Results for the PPI analysis. The left lateral occipitotemporal (LOTC) was used as the source region. A cluster in the left Intraparietal sulcus (IPS) showed significantly higher connectivity with the left LOTC in patients than in healthy volunteers (A). A voxel-level threshold of *p* < 0.001 (uncorrected) and a cluster-level threshold of *p* < 0.05 (family-wise error corrected) was applied. The colour bar represents *t* values. This left IPS cluster was involved in a region-of-interest-based analysis to associate its values with patients’ scores in the meaningless gesture imitation and gesture production tasks performed outside the scanner. Multiple regression models were applied to the patient sample, involving age and total lesion size as covariates.

Further PPIs investigating group-related connectivity changes with the right LOTC or either the right or the left and right EBA ROIs for ‘Bodies’ revealed no significant results.

We carried out a follow up regression analysis to see if this PPI connectivity result between left LOTC and IPS was related to meaningless gesture imitation apraxia scores in patients. The left IPS cluster obtained from the whole-brain PPI analysis (Figure 4A) was used as a ROI, with its mean values from the PPI analysis extracted for each patient. This analysis revealed a significant association with the meaningless gesture imitation scores (*β* = −0.46, *t* = −2.25, *p* = 0.035), controlling for age and total lesion size (Figure 4B). This indicates that higher connectivity was associated with lower test scores, i.e., more impairment in meaningless gesture imitation. However, the association with gesture production was not significant (*β* = −0.20, *t* = −0.92, *p* = 0.37). We repeated this analysis by further excluding two patients who had lesions in the left LOTC or IPS (24 patients remaining). The association with the gesture imitation scores remained significant (*β* = −0.53, *t* = −2.40, *p* = 0.025).

## Discussion

In this study we investigated the neural correlates underlying the perception of Tools and Bodies in left hemisphere stroke patients comparing them to healthy participants, using a localiser task which is known to activate a reliable network of brain regions centred within and connected to the lateral occipito-temporal cortex (LOTC) (Gallivan et al., 2013).

Over recent years, LOTC has been increasingly recognized for representing various aspects of action, ranging from the perception of tools and bodies to their typical movements (Mahon and Almeida, 2024). These representations are critical for understanding and performing actions. It is thought that activity in LOTC forms representational spaces that capture perceptual, semantic, and motor dimensions of actions, reflecting how actions alter the state of the world (Lingnau and Downing, 2015; Chen et al., 2017; Mahon and Almeida, 2024).

Our findings support recent findings suggesting an important role for the LOTC in limb apraxia following stroke (Sperber et al., 2019; Pizzamiglio et al., 2019), with two key outcomes: 1) reduced selectivity in the contralateral LOTC, and 2) increased connectivity between LOTC and the left intraparietal sulcus (L-IPS). We discuss these in the sections below.

### 1) Reduced selectivity in contralesional Right LOTC & EBA

Our study found comparable levels of activation in the lateral occipito-temporal cortex (LOTC) and extrastriate body area (EBA) during the tools and bodies localizers across stroke patients and healthy volunteers. However representational similarity analysis (RSA) revealed subtle differences in neural representations. Specifically, dissimilarity measures for tools were reduced in the contralesional (right) hemisphere in patients, suggesting altered neural tuning, despite similar global activation patterns across groups. A similar trend was observed for bodies in the EBA, indicating that these changes may reflect ‘functional diaschisis,’ where distant lesions disrupt functional networks (Carrera and Tononi, 2014).

These findings parallel previous studies on cognitive recovery, such as in language after stroke, which showed preserved activation in temporal lobe areas despite changes in behavioural outcomes due to altered neural tuning (Leff et al., 2002; Crinion et al., 2006). Thus, absence of significant changes in fMRI activation does not preclude neural-level alterations.

It is noteworthy that the fMRI tuning differences observed when comparing patients and healthy volunteers were in the contra-lesional, non-affected hemisphere. Previous studies have highlighted the role of the opposite hemisphere in the recovery of visuo-perceptual abilities post-stroke and its influence on motor recovery ((Karolis et al., 2019;Mattos et al., 2021).

This study highlights the value of task-related fMRI in detecting nuanced changes in brain organization, particularly during the chronic post-stroke stage. These methods can elucidate how perceptual functions adapt through displaced or retuned processing to maintain stable behavioural performance (Stefaniak et al., 2020).

### 2) Increased functional connectivity between LOTC and IPS within left hemisphere

Our PPI analysis revealed increased functional connectivity between the LOTC and the left intraparietal sulcus (L-IPS) when patients viewed tools, compared to healthy controls.

The L-IPS plays a critical role in visuomotor representations that guide actions and is associated with the transport phase of reaching to grasp tasks (Rossit et al., 2013; Gallivan et al., 2015). Its activation is often linked to task difficulty in visuospatial and semantic tasks (Humphreys and Lambon Ralph, 2017; Humphreys et al., 2022). This area works together with the lateral occipito-temporal cortex (LOTC), which is involved in the conceptual processing of actions and events, as well as the recognition of objects—especially tools—that are frequently either the agents or targets of actions (Brambati et al., 2006; Kalénine et al., 2010; Mahon et al., 2007; Buxbaum et al., 2014; Lingnau and Downing, 2015; Tranel et al., 2003; Wurm et al., 2017; Wurm and Caramazza, 2022).

This increased connectivity between L-IPS and LOTC was associated with poorer performance in the meaningless gesture imitation task, a known sensitive measure of apraxia (Bickerton et al., 2012). These areas have been implicated in semantic control (Humphreys et al., 2022), whilst meaningless gesture imitation has been associated with higher order functions including selecting between postures (Rounis et al., 2021) with harder ones being more difficult for patients to solve (Achilles et al., 2016). Interestingly, our localizer task involved both manipulable and non-manipulable objects, and the former may overlap with body representations in LOTC, especially when participants viewed upper limb areas. LOTC is known to represent hand postures and motor properties of objects (Bracci et al., 2018; Wurm et al., 2017). This enhanced connectivity may reflect a compensatory mechanism, where the brain recruits additional visuomotor resources from dorsal stream areas to support action recognition and performance (Zimmermann et al., 2018; Zhang et al., 2021; Mahon and Almeida, 2024).

#### Limitations

We note that the sample size for our imaging analyses (N=27 had actual scans and 26 were analysed as 1 patient also performed below 3SD on the 1-back task) and inclusion of left hemisphere limits the conclusions we can draw from our results. Nevertheless, the results presented here have survived small volume correction even when removing patients in whom lesions affected the seed or results sites. Future studies would be helpful to explore further the influence of apraxia subtypes as well as lesioned hemisphere (therefore including right hemisphere lesions) on neural tuning and connectivity in larger patient populations.

### Implications for the Role of Parietal Lobe Damage in Apraxia

Our findings underscore the role of the left parietal lobe, particularly the intraparietal sulcus, in meaningless gesture imitation, a task more sensitive to apraxic deficits than tool-use pantomime ((Goldenberg, 2009; Achilles et al., 2016). Both animal and human studies have shown that the parietal cortex plays a role in encoding spatial relationships between body parts and objects (Arbib et al., 2009; Orban and Caruana, 2014; Osiurak and Badets, 2016; Osiurak et al., 2021). This area is crucial for maintaining a dynamic representation of body schema, which continuously updates the body’s spatial configuration, particularly during action planning (Haggard P, Wolpert DM., 2005; (Gallagher and Brøsted Sørensen, 2006; Zimmermann et al., 2012).

Goodale and Milner’s (1992) perception-action model emphasizes the distinct roles of the dorsal and ventral streams in guiding goal-directed movements. The dorsal occipito-parietal stream processes visuospatial information for goal-directed actions, while the ventral occipito-temporal stream is specialized for object recognition and identification. In this framework, the LOTC and EBA are key to perceiving manipulable objects and body parts, respectively (Downing & Lignau, 2015; Zimmermann et al., 2018). Together with the parietal lobe that represents body state estimation(Shadmehr and Krakauer, 2008), this network integrates motor planning with representations of the desired postural configurations for future actions (Astafiev et al., 2004; Zimmermann et al., 2012; Zhang et al., 2021). This ability to anticipate future states is disrupted in patients with limb apraxia ((Buxbaum et al., 2014; Sperber et al., 2019; Pizzamiglio et al., 2019a; Pizzamiglio et al., 2019b; Vigliocco et al., 2020).

## Conclusion

In conclusion our findings highlight complex disruptions in both perceptual and motor networks in limb apraxia at the chronic stages following a stroke. Changes in LOTC selectivity and its enhanced connectivity with L-IPS point to altered functional dynamics within these networks. These insights provide a deeper understanding of how perceptual and motor impairments interact in apraxia and suggest that targeting these disrupted networks through neurorehabilitation may improve functional outcomes.

## Data Availability

All data (except from video recordings of patients, which are identifiable) produced in the present study are available upon reasonable request to the authors

## Acknowledgements

We would like to thank the participants taking part in this study. This study was supported by grants from the British Medical Association and Oxford University Graduate Programme for Clinical Lecturers to Dr Elisabeth Rounis, who also currently holds a UKRI Clinical Academic Partnership with Professor Lambon Ralph and Dr Ajay Halai.

## References

Achilles, E.I.S., Fink, G.R., Fischer, M.H., Dovern, A., Held, A., Timpert, D.C., Schroeter, C., Schuetz, K., Kloetzsch, C., Weiss, P.H., 2016. Effect of meaning on apraxic finger imitation deficits. Neuropsychologia 82, 74–83. 10.1016/j.neuropsychologia.2015.12.022

Ant, J.M., Niessen, E., Achilles, E.I.S., Saliger, J., Karbe, H., Weiss, P.H., Fink, G.R., 2019. Anodal tDCS over left parietal cortex expedites recovery from stroke-induced apraxic imitation deficits: a pilot study. Neurol Res Pract 1, 38. 10.1186/s42466-019-0042-0

Arbib, M.A., Bonaiuto, J.B., Jacobs, S., Frey, S.H., 2009. Tool use and the distalization of the end-effector. Psychol Res 73, 441–462. 10.1007/s00426-009-0242-2

Astafiev, S.V., Stanley, C.M., Shulman, G.L., Corbetta, M., 2004. Extrastriate body area in human occipital cortex responds to the performance of motor actions. Nat Neurosci 7, 542–548. 10.1038/nn1241

Bickerton, W.-L., Riddoch, M.J., Samson, D., Balani, A.B., Mistry, B., Humphreys, G.W., 2012. Systematic assessment of apraxia and functional predictions from the Birmingham Cognitive Screen. J Neurol Neurosurg Psychiatry 83, 513–521. 10.1136/jnnp-2011-300968

Binkofski, F., Buxbaum, L.J., 2013. Two action systems in the human brain. Brain Lang 127, 222–229. 10.1016/j.bandl.2012.07.007

Bracci, S., Caramazza, A., Peelen, M.V., 2018. View-invariant representation of hand postures in the human lateral occipitotemporal cortex. Neuroimage 181, 446–452. 10.1016/j.neuroimage.2018.07.001

Brambati, S.M., Myers, D., Wilson, A., Rankin, K.P., Allison, S.C., Rosen, H.J., Miller, B.L., Gorno-Tempini, M.L., 2006. The anatomy of category-specific object naming in neurodegenerative diseases. J Cogn Neurosci 18, 1644–1653. 10.1162/jocn.2006.18.10.1644

Buxbaum, L.J., Randerath, J., 2018. Limb apraxia and the left parietal lobe. Handb Clin Neurol 151, 349–363. 10.1016/B978-0-444-63622-5.00017-6

Buxbaum, L.J., Shapiro, A.D., Coslett, H.B., 2014. Critical brain regions for tool-related and imitative actions: a componential analysis. Brain 137, 1971–1985. 10.1093/brain/awu111

Carrera, E., Tononi, G., 2014. Diaschisis: past, present, future. Brain 137, 2408–2422. 10.1093/brain/awu101

Catani, M., ffytche, D.H., 2005. The rises and falls of disconnection syndromes. Brain 128, 2224–2239. 10.1093/brain/awh622

Chen, L., Lambon Ralph, M.A., Rogers, T.T., 2017. A unified model of human semantic knowledge and its disorders. Nat Hum Behav 1, 0039. 10.1038/s41562-016-0039

Cloutman, L.L., 2013. Interaction between dorsal and ventral processing streams: where, when and how? Brain Lang 127, 251–263. 10.1016/j.bandl.2012.08.003

Crinion, J.T., Warburton, E.A., Lambon-Ralph, M.A., Howard, D., Wise, R.J.S., 2006. Listening to narrative speech after aphasic stroke: the role of the left anterior temporal lobe. Cereb Cortex 16, 1116–1125. 10.1093/cercor/bhj053

Esteban, O., Markiewicz, C.J., Blair, R.W., Moodie, C.A., Isik, A.I., Erramuzpe, A., Kent, J.D., Goncalves, M., DuPre, E., Snyder, M., Oya, H., Ghosh, S.S., Wright, J., Durnez, J., Poldrack, R.A., Gorgolewski, K.J., 2019. fMRIPrep: a robust preprocessing pipeline for functional MRI. Nat Methods 16, 111–116. 10.1038/s41592-018-0235-4

Gallagher, S., Brøsted Sørensen, J., 2006. Experimenting with phenomenology. Conscious Cogn 15, 119–134. 10.1016/j.concog.2005.03.002

Gallivan, J.P., Barton, K.S., Chapman, C.S., Wolpert, D.M., Flanagan, J.R., 2015. Action plan co-optimization reveals the parallel encoding of competing reach movements. Nat Commun 6, 7428. 10.1038/ncomms8428

Gallivan, J.P., McLean, D.A., Valyear, K.F., Culham, J.C., 2013. Decoding the neural mechanisms of human tool use. eLife 2, e00425. 10.7554/eLife.00425

Goldenberg, G., 2009. Apraxia and the parietal lobes. Neuropsychologia 47, 1449–1459. 10.1016/j.neuropsychologia.2008.07.014

Goldenberg, G., Karnath, H.-O., 2006. The neural basis of imitation is body part specific. J Neurosci 26, 6282–6287. 10.1523/JNEUROSCI.0638-06.2006

Goodale, M.A., Milner, A.D., 1992. Separate visual pathways for perception and action. Trends Neurosci 15, 20–25. 10.1016/0166-2236(92)90344-8

Gorgolewski, K., Burns, C.D., Madison, C., Clark, D., Halchenko, Y.O., Waskom, M.L., Ghosh, S.S., 2011. Nipype: a flexible, lightweight and extensible neuroimaging data processing framework in python. Front Neuroinform 5, 13. 10.3389/fninf.2011.00013

Haggard P, Wolpert DM., 2005. Disorders of Body Scheme, in: Higher-Order Motor Disorders. Oxford University Press.

Hebart, M.N., Görgen, K., Haynes, J.-D., 2014. The Decoding Toolbox (TDT): a versatile software package for multivariate analyses of functional imaging data. Front Neuroinform 8, 88. 10.3389/fninf.2014.00088

Heilman, K.M., Watson, R.T., 2008. The disconnection apraxias. Cortex 44, 975–982. 10.1016/j.cortex.2007.10.010

Hoeren, M., Kümmerer, D., Bormann, T., Beume, L., Ludwig, V.M., Vry, M.-S., Mader, I., Rijntjes, M., Kaller, C.P., Weiller, C., 2014. Neural bases of imitation and pantomime in acute stroke patients: distinct streams for praxis. Brain 137, 2796–2810. 10.1093/brain/awu203

Humphreys, G.F., Jung, J., Lambon Ralph, M.A., 2022. The convergence and divergence of episodic and semantic functions across lateral parietal cortex. Cereb Cortex 32, 5664–5681. 10.1093/cercor/bhac044

Humphreys, G.F., Lambon Ralph, M.A., 2017. Mapping Domain-Selective and Counterpointed Domain-General Higher Cognitive Functions in the Lateral Parietal Cortex: Evidence from fMRI Comparisons of Difficulty-Varying Semantic Versus Visuo-Spatial Tasks, and Functional Connectivity Analyses. Cereb Cortex 27, 4199–4212. 10.1093/cercor/bhx107

Kalénine, S., Buxbaum, L.J., Coslett, H.B., 2010. Critical brain regions for action recognition: lesion symptom mapping in left hemisphere stroke. Brain 133, 3269–3280. 10.1093/brain/awq210

Karolis, V.R., Corbetta, M., Thiebaut de Schotten, M., 2019. The architecture of functional lateralisation and its relationship to callosal connectivity in the human brain. Nat Commun 10, 1417. 10.1038/s41467-019-09344-1

Kriegeskorte, N., Mur, M., Bandettini, P., 2008. Representational similarity analysis - connecting the branches of systems neuroscience. Front Syst Neurosci 2, 4. 10.3389/neuro.06.004.2008

Leff, A., Crinion, J., Scott, S., Turkheimer, F., Howard, D., Wise, R., 2002. A physiological change in the homotopic cortex following left posterior temporal lobe infarction. Ann Neurol 51, 553–558. 10.1002/ana.10181

Lingnau, A., Downing, P.E., 2015. The lateral occipitotemporal cortex in action. Trends Cogn Sci 19, 268–277. 10.1016/j.tics.2015.03.006

Mahon, B.Z., Almeida, J., 2024. Reciprocal interactions among parietal and occipito-temporal representations support everyday object-directed actions. Neuropsychologia 198, 108841. 10.1016/j.neuropsychologia.2024.108841

Mahon, B.Z., Milleville, S.C., Negri, G.A.L., Rumiati, R.I., Caramazza, A., Martin, A., 2007. Action-related properties shape object representations in the ventral stream. Neuron 55, 507–520. 10.1016/j.neuron.2007.07.011

Mattos, D.J.S., Rutlin, J., Hong, X., Zinn, K., Shimony, J.S., Carter, A.R., 2021. White matter integrity of contralesional and transcallosal tracts may predict response to upper limb task-specific training in chronic stroke. Neuroimage Clin 31, 102710. 10.1016/j.nicl.2021.102710

Orban, G.A., Caruana, F., 2014. The neural basis of human tool use. Front Psychol 5, 310. 10.3389/fpsyg.2014.00310

Osiurak, F., Badets, A., 2016. Tool use and affordance: Manipulation-based versus reasoning-based approaches. Psychol Rev 123, 534–568. 10.1037/rev0000027

Osiurak, F., Reynaud, E., Baumard, J., Rossetti, Y., Bartolo, A., Lesourd, M., 2021. Pantomime of tool use: looking beyond apraxia. Brain Commun 3, fcab263. 10.1093/braincomms/fcab263

Pazzaglia, M., Smania, N., Corato, E., Aglioti, S.M., 2008. Neural underpinnings of gesture discrimination in patients with limb apraxia. J. Neurosci. 28, 3030–3041. 10.1523/JNEUROSCI.5748-07.2008

Pizzamiglio, G., Zhang, Z., Duta, M., Rounis, E., 2019a. Factors Influencing Manipulation of a Familiar Object in Patients With Limb Apraxia After Stroke. Front Hum Neurosci 13, 465. 10.3389/fnhum.2019.00465

Pizzamiglio, G., Zhang, Z., Kolasinski, J., Riddoch, J.M., Passingham, R.E., Mantini, D., Rounis, E., 2019b. A Role for the Action Observation Network in Apraxia After Stroke. Front Hum Neurosci 13, 422. 10.3389/fnhum.2019.00422

Ramayya, A.G., Glasser, M.F., Rilling, J.K., 2010. A DTI investigation of neural substrates supporting tool use. Cereb Cortex 20, 507–516. 10.1093/cercor/bhp141

Rossit, S., McAdam, T., McLean, D.A., Goodale, M.A., Culham, J.C., 2013. fMRI reveals a lower visual field preference for hand actions in human superior parieto-occipital cortex (SPOC) and precuneus. Cortex 49, 2525–2541. 10.1016/j.cortex.2012.12.014

Rounis, E., Banca, P., Voon, V., 2016. Deficits in Limb Praxis in Patients With Obsessive-Compulsive Disorder. J Neuropsychiatry Clin Neurosci 28, 232–235. 10.1176/appi.neuropsych.15090233

Rounis, E., Binkofski, F., 2023. Limb Apraxias: The Influence of Higher Order Perceptual and Semantic Deficits in Motor Recovery After Stroke. Stroke 54, 30–43. 10.1161/STROKEAHA.122.037948

Rounis, E., Halai, A., Pizzamiglio, G., Lambon Ralph, M.A., 2021. Characterising factors underlying praxis deficits in chronic left hemisphere stroke patients. Cortex 142, 154–168. 10.1016/j.cortex.2021.04.019

Rounis, E., Thompson, E., Scandola, M., Nozais, V., Pizzamiglio, G., de Schotten, M.T., Pacella, V., 2024. A preliminary study of white matter disconnections underlying deficits in praxis in left hemisphere stroke patients. Brain Struct Funct. 10.1007/s00429-024-02814-3

Schmidt, C.C., Achilles, E.I.S., Fink, G.R., Weiss, P.H., 2022. Distinct cognitive components and their neural substrates underlying praxis and language deficits following left hemisphere stroke. Cortex 146, 200–215. 10.1016/j.cortex.2021.11.004

Shadmehr, R., Krakauer, J.W., 2008. A computational neuroanatomy for motor control. Exp Brain Res 185, 359–381. 10.1007/s00221-008-1280-5

Sperber, C., Wiesen, D., Goldenberg, G., Karnath, H.-O., 2019. A network underlying human higher-order motor control: Insights from machine learning-based lesion-behaviour mapping in apraxia of pantomime. Cortex 121, 308–321. 10.1016/j.cortex.2019.08.023

Stefaniak, J.D., Halai, A.D., Lambon Ralph, M.A., 2020. The neural and neurocomputational bases of recovery from post-stroke aphasia. Nat Rev Neurol 16, 43–55. 10.1038/s41582-019-0282-1

Tranel, D., Kemmerer, D., Adolphs, R., Damasio, H., Damasio, A.R., 2003. Neural correlates of conceptual knowledge for actions. Cogn Neuropsychol 20, 409–432. 10.1080/02643290244000248

Umarova, R.M., Saur, D., Schnell, S., Kaller, C.P., Vry, M.-S., Glauche, V., Rijntjes, M., Hennig, J., Kiselev, V., Weiller, C., 2010. Structural connectivity for visuospatial attention: significance of ventral pathways. Cereb Cortex 20, 121–129. 10.1093/cercor/bhp086

Vigliocco, G., Krason, A., Stoll, H., Monti, A., Buxbaum, L.J., 2020. Multimodal comprehension in left hemisphere stroke patients. Cortex 133, 309–327. 10.1016/j.cortex.2020.09.025

Weiller, C., Bormann, T., Saur, D., Musso, M., Rijntjes, M., 2011. How the ventral pathway got lost: and what its recovery might mean. Brain Lang 118, 29–39. 10.1016/j.bandl.2011.01.005

Wurm, M.F., Caramazza, A., 2022. Two “what” pathways for action and object recognition. Trends Cogn Sci 26, 103–116. 10.1016/j.tics.2021.10.003

Wurm, M.F., Caramazza, A., Lingnau, A., 2017. Action Categories in Lateral Occipitotemporal Cortex Are Organized Along Sociality and Transitivity. J Neurosci 37, 562–575. 10.1523/JNEUROSCI.1717-16.2016

Zhang, L., Pini, L., Kim, D., Shulman, G.L., Corbetta, M., 2023. Spontaneous Activity Patterns in Human Attention Networks Code for Hand Movements. J Neurosci 43, 1976–1986. 10.1523/JNEUROSCI.1601-22.2023

Zhang, Z., Zeidman, P., Nelissen, N., Filippini, N., Diedrichsen, J., Bracci, S., Friston, K., Rounis, E., 2021. Neural Correlates of Hand-Object Congruency Effects during Action Planning. J Cogn Neurosci 33, 1487–1503. 10.1162/jocn_a_01728

Zimmermann, M., Mars, R.B., de Lange, F.P., Toni, I., Verhagen, L., 2018. Is the extrastriate body area part of the dorsal visuomotor stream? Brain Struct Funct 223, 31–46. 10.1007/s00429-017-1469-0

Zimmermann, M., Meulenbroek, R.G.J., de Lange, F.P., 2012. Motor planning is facilitated by adopting an action’s goal posture: an fMRI study. Cereb Cortex 22, 122–131. 10.1093/cercor/bhr098

